# Comparison of preprints and final journal publications from COVID-19 Studies: Discrepancies in results reporting and spin in interpretation

**DOI:** 10.1101/2021.04.12.21255329

**Authors:** Lisa Bero, Rosa Lawrence, Louis Leslie, Kellia Chiu, Sally McDonald, Matthew J Page, Quinn Grundy, Lisa Parker, Stephanie L Boughton, Jamie J Kirkham, Robin Featherstone

## Abstract

**Objective:** To compare results reporting and the presence of spin in COVID-19 study preprints with their finalized journal publications

**Design:** Cross-sectional

**Setting:** International medical literature

**Participants:** Preprints and final journal publications of 67 interventional and observational studies of COVID-19 treatment or prevention from the Cochrane COVID-19 Study Register published between March 1, 2020 and October 30, 2020

**Main outcome measures:** Study characteristics and discrepancies in 1) Results reporting (number of outcomes, outcome descriptor, measure (e.g., PCR test), metric (e.g., mean change from baseline), assessment time point (e.g., 1 week post treatment), data reported (e.g., effect estimate and measures of precision), reported statistical significance of result, type of statistical analysis (e.g., chi-squared test), subgroup analyses (if any), whether outcome was identified as primary or secondary and 2) Spin (reporting practices that distort the interpretation of results so that results are viewed more favorably).

**Results:** Of 67 included studies, 23 (34%) had no discrepancies in results reporting between preprints and journal publications. Fifteen (22%) studies had at least one outcome that was included in the journal publication, but not the preprint; 8 (12%) had at least one outcome that was reported in the preprint only. For outcomes that were reported in both preprints and journals, common discrepancies were differences in numerical values and statistical significance, additional statistical tests and subgroup analyses conducted in journal publications, and longer follow-up times for outcome assessment in journal publications.

At least one instance of spin occurred in both preprints and journals in 23 / 67 (34%) studies, the preprint only in 5 (7%) studies, and the journal publications only in 2 (3%) of studies. Spin was removed between the preprint and journal publication in 5/67 (7%) studies; but added in 1/67 (1%) study.

**Conclusions:** The COVID-19 preprints and their subsequent journal publications were largely similar in reporting of study characteristics, outcomes and spin. All COVID-19 studies published as preprints and journal publications should be critically evaluated for discrepancies and spin.

**EQUATOR REPORTING GUIDELINE:** STROBE

**What is already known on this topic:**

- Selective and incomplete reporting of results and spin are threats to the trustworthiness and validity of research.
- These reporting practices could be particularly dangerous for users of COVID-19 research as they can inflate the efficacy of interventions and underestimate harms.
- Given the high prevalence, visibility, and potentially rapid implementation of COVID-19 research published as preprints, it is important to compare components of results reporting and the presence of spin in COVID-19 studies on treatment or prevention that are published both as preprints and journal publications.

**What this study adds:**

- This comparison of 67 COVID-19 preprints related to treatment or prevention and their subsequent journal publications found they were largely similar in reporting of study characteristics, components of results reporting and spin in interpretation.
- Even a few important discrepancies could impact decision making.

## INTRODUCTION

Preprints have been advocated as a means for rapid sharing and updating of research findings, which could be particularly valuable during a pandemic.[1] Preprints are non-peer-reviewed postings of research articles. Preprints have been a common form of publication in the natural sciences for decades, and more recently in the life sciences. In 2019, BMJ, Yale and Cold Spring Harbor Laboratory launched medRxiv, a preprint server dedicated to clinical and health sciences research.

In April 2020, medRxiv published between 50 and 100 COVID-19-related preprints daily.[1] The accelerated pace of research related to COVID-19 has increased the potential impact and risk of using preprints. Widespread public dissemination of preprints may spread misinformation.[2] A study comparing 34 preprints and 62 publications about therapies for COVID-19 found that publications had significantly more citations than the preprints (median of 22 vs 5.5 citations; *P*⍰=⍰.01), but there were no significant differences for attention and online engagement metrics.[3]

Most preprint servers conduct some type of screening prior to posting, commonly related to the scope of the article, plagiarism, and compliance with legal and ethical requirements[4], but preprints have not been peer-reviewed and may not meet the methodological and reporting requirements of a journal. A review of the medRxiv preprint server one year after its launch found that 9967 of 11164 (89%) of submissions passed screening.[5] It is not clear whether or how preprint servers might screen for quality of results reporting or spin.[6,7] Spin refers to specific reporting practices that distort the interpretation of results so that results are viewed more favorably.

Preliminary studies suggest that reporting discrepancies may exist between preprints and subsequent publications. However, there has been no systematic assessments of results reporting or spin between preprints and their final journal publications. Carneiro et al.counted reported items from a checklist meant to cover common points from multiple reporting guidelines and found reporting to be a little higher in journal articles, both in a set of bioRxiv preprints matched to their journal publication (n=56 article/group) and in an unmatched set (n=76 articles/group).[8] An analysis of preprints from arXiv, a primarily physics/ mathematics preprint server, and their journal publications using text comparison algorithms found little difference between preprints and published articles.[9] However, an analysis of medRxiv and bioRxiv preprints related to COVID-19 pharmacological interventions found that only 24% (23/97) of preprints were published in a journal within 0 to 98 days (median: 42.0 days). Among these, almost half (11/23, 48%) had modifications in the title or results section, although the nature of these modifications is not described.[10] An analysis of spin in preprints and journal publications for COVID-19 trials found a single difference between 2 matched pairs preprint and their journal publications: the discussion of limitations in the abstract. Limitations were discussed in the abstract of one article, but not its accompanying preprint. [11] An analysis of 66 preprint-article pairs of COVID-19 studies found 38% had changes in study results, such as a numeric change in hazard ratio or a change in p value, and 29% had changes in abstract conclusions, most commonly from positive without reporting uncertainty in the preprint to positive with reporting of uncertainty in the article.[12]

The trustworthiness and validity of scientific publications, even after peer review, are weakened by a variety of problems.[13,14] Selective and incomplete results reporting[15,16] and spin[17,18] are two critical threats, especially for clinical studies of treatment or prevention. These reporting practices could be particularly dangerous for users of COVID-19 research as they can inflate the efficacy of interventions and underestimate harms. Given the high prevalence, visibility, and potentially rapid implementation of COVID-19 research published as preprints, this study is the first to compare components of outcome reporting and the presence of spin in COVID-19 studies on treatment or prevention that are published both as preprints and journal publications.

## METHODS

The protocol for this study was registered in the Open Science Framework.[19]

### Data Source and Search Strategy

We sampled studies from the Cochrane COVID-19 Study Register (https://covid-19.cochrane.org/), a freely-available, continually-updated, annotated reference collection of human primary studies on COVID-19, including interventional, observational, diagnostic, prognostic, epidemiological and qualitative designs. The register is “study-based,” meaning references to the same study (e.g., press releases, trial registry records, preprints, journal pre-proofs, journal final publications, retraction notices) are all linked to a single study identifier. References are screened for eligibility to determine if they are primary studies (e.g., not opinion pieces or narrative reviews). Data sources for the Cochrane COVID-19 Study Register at the time of the search included ClinicalTrials.gov, the International Clinical Trials Registry Platform (ICTRP), PubMed, medRxiv and Embase.com. The Cochrane register prioritizes medRxiv as a preprint source as an internal sensitivity analysis in May 2020 showed that 90% (166/185) of the preprints that were eligible for systematic reviews came from this source. The register also includes preprints records sourced from PubMed.

All studies in the register are classified by study design (interventional, observational, modelling, qualitative, other or unclear) and research aim (prevention, treatment and management, diagnostic/prognostic, epidemiology, health services research, mechanism, transmission, other). Studies may be classified as having multiple research aims. Four searches using the register’s search filters for study reference types (preprints and journal articles) and study characteristics (study type and study aim) were used to retrieve references with a study aim of a) treatment and management or b) prevention and classified as interventional or observational (see OSF project for the complete search strategies: (https://osf.io/5ru8w/?view_only=fe509bf54c104354a1e12f011bdff66a). As the register is updated daily, we repeated the search. The Cochrane COVID-19 Study Register was first searched by RF on October 13, and updated on October 29, 2020. Results were exported to Excel and duplicates manually identified. The searches identified 297 references for 117 studies, with 67 (21 interventional, 46 observational) that met our inclusion and exclusion criteria for study selection (Figure 1).

**Figure 1.**
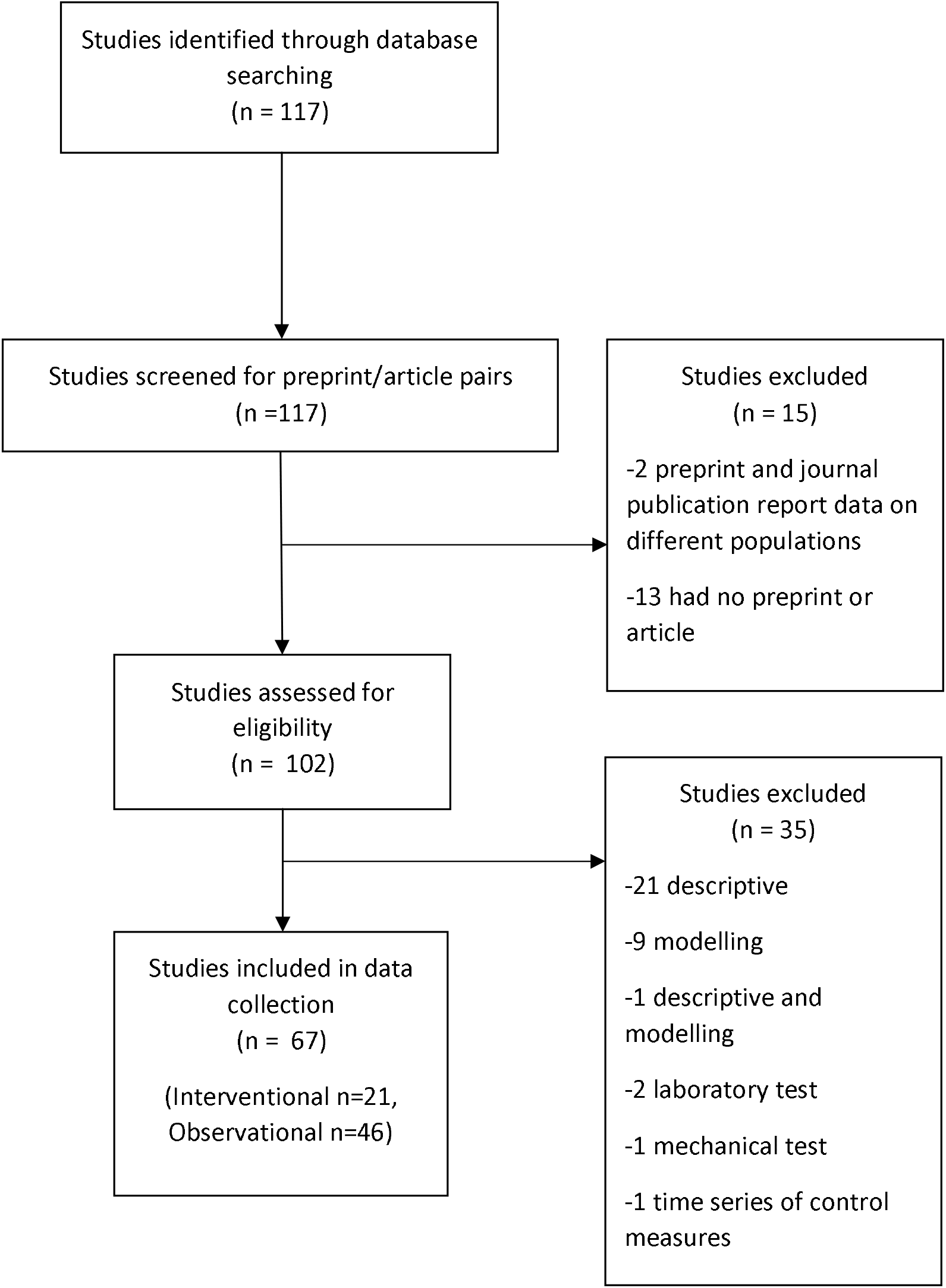
Flowchart of study inclusion.

### Inclusion and exclusion criteria for study selection

We included studies of COVID-19 treatment or prevention identified in the search that had both a posted preprint and final journal publication.

We included studies with aims of diagnosis/prognosis, epidemiology, health services research, mechanism, transmission and other if they also had an aim coded as a) treatment and management or b) prevention. We excluded modelling studies, qualitative studies and studies that reported only descriptive data (e.g., demographic characteristics).

We screened all records for each included study to identify posted preprints and journal publications from each study. We excluded duplicates and records for protocols, trial registries, commentaries, letters to the editor, news articles, and press releases. We excluded records that did not report results and non-English records.

We compared the preprint and journal publication for each included study. In the case of multiple preprints or journal publications reporting study results, we selected the first preprint version and the final journal publication that reported on similar study populations. This was to ensure that the preprint version evaluated in our study had not been altered in response to any comments, which could constitute a form of peer review, and that it was representative of the version most likely to be seen by clinicians, journalists and other research users as new research became available.

### Data extraction

Ten investigators (LB, SLB, KC, QG, JJK, LL, RL, SMc, LP, MJP) working independently in pairs extracted data from the included studies. Discrepancies in data extraction were resolved by consensus. If agreement could not be reached, an investigator who was not part of the coding pair resolved the discrepancies. All extracted data from the included studies was stored in REDCap, a secure web-based application for the collection and management of data.[20] We extracted data from the medRxiv page and PDF for preprints and the online publication or PDF for journal articles. We extracted data on results reporting, presence of spin and study characteristics as described below.

### Study characteristics

For each preprint, we recorded the earliest posting date; for each journal publication we extracted the submitted/received, reviewed, revised, accepted and published date(s), where available.

From each journal publication, we extracted: authors, title, funding source, author conflicts of interests, ethics approval, country of study, and sample size. For the accompanying preprint, we determined if these study characteristics were also reported. If they were, and the content of the item differed between the preprint and publication, details of the discrepancy were recorded. In addition, we recorded discrepancies between the preprint and journal publication in demographic characteristics of study participants (e.g., sex, race/ethnicity, diagnosis), discussion of limitations (regardless of whether there was a labeled limitations section or not), and tables and figures.

### Primary outcomes

Our primary outcome measures were 1) discrepancies in results reporting between preprints and journal publications and 2) presence and type of spin in preprints and journal publications.

### Results reporting

We collected data on discrepancies in 1) number of outcomes reported in preprints and journal publications and, for outcomes reported in both preprints and journal publications, 2) components of results reporting. For each journal publication and preprint, we recorded the number of outcomes reported and, whether outcomes were reported only in the preprint or journal publication, and the outcome descriptor (e.g., mortality, hospitalization, transmission, immunogenicity, harms).

For outcomes that were reported in both preprints and journal publications,, we collected data on components of outcome reporting based on recommendations for clinical study results reporting.[16,21] We recorded whether there were discrepancies between any components of outcome reporting between journal publications and preprints. We extracted the text relevant to each discrepancy:

⍰ Measure (e.g., PCR test)
⍰ Metric (e.g., mean change from baseline, proportion of people)
⍰ Time point at which the assessment was made (e.g., 1 week after starting treatment).
⍰ Numerical values reported (e.g., effect estimate and measure of precision)
⍰ Statistical significance of result (as reported)
⍰ Type of statistical analysis (e.g., regression, chi-squared test)
⍰ Subgroup analyses (if any)
⍰ Whether outcome was identified as primary or secondary

### Spin

Studies have used a variety of methods to measure spin in randomized controlled trials and observational studies.[17] Based on our previously developed typology of spin derived from a systematic review of spin studies,[17] we developed and pretested a coding tool for spin that can be applied to both interventional and observational studies of treatment or prevention. In the context of research on treatment or prevention of COVID-19, the most meaningful consequences of spin are overinterpretation of efficacy and underestimation of harms. Therefore, our tool emphasizes these manifestations of spin. We searched the abstracts and full text of each preprint and journal publication for 3 primary categories of spin, and accompanying subcategories:

1. Inappropriate interpretation given study design
  ⍰ Claiming causality in non-randomized studies
  ⍰ Interpreting a lack of statistical significance as equivalence
  ⍰ Interpreting a lack of statistical significance of harm measures as safety
  ⍰ Claim of any significant difference despite lack of statistical test
  ⍰ Other
2. Inappropriate extrapolations or recommendations
  ⍰ Suggestion that the intervention or exposure is more clinically relevant or useful than is justified given the study design
  ⍰ Recommendation made to population groups / contexts outside of those investigated
  ⍰ (Observational) Expressing confidence in an intervention or exposure without suggesting the need for further confirmatory studies
  ⍰ Other
3. Selectively focusing on positive results or more favorable data presentation
  ⍰ Discussing only significant (non-primary) results to distract from non-significant primary results
  ⍰ Omitting non-significant results from abstract / discussion / conclusion
  ⍰ Claiming significant effects for non-significant results
  ⍰ Acknowledge statistically nonsignificant results from the primary outcome but emphasize the beneficial effect of treatment
  ⍰ Describing non-significant results as “trending towards significance”
  ⍰ Mentioning adverse effects in the abstract / discussion /conclusion but minimizing their potential effect or importance
  ⍰ Misleading description of study design as one that is more robust
  ⍰ Use of linguistic spin
  ⍰ Other

## Analysis

We report the frequency and types of discrepancies in study characteristics and results reporting between preprints and journal publications. We report the proportion of preprints and journal publications with spin and the types of spin. We iteratively analyzed the text descriptions of discrepancies identified; we grouped descriptions into common categories, while still accounting for all instances of discrepant reporting, even if it only occurred once, to demonstrate the range of the phenomenon.

To determine whether preprints that were posted after an article received peer review influenced the number of discrepancies, we conducted a *post hoc* sensitivity analysis by removing 7 studies where the preprint was posted up to 7 days before the revision, acceptance, or publication dates of the journal publication.

Our protocol modification, list of included preprints and journal publications, data dictionary and dataset are available in our OSF project linked to our protocol: https://osf.io/5ru8w/?view_only=fe509bf54c104354a1e12f011bdff66a.

## Ethics approval

This study analyzes publicly available information and is exempt from ethics review.

This study had no patient or public involvement.

## RESULTS

### Study characteristics

Of the 67 included studies, 57 were studies of treatment and management, 9 of prevention, and 1 of both. The preprints and journal publications were published between March 1, 2020 and October 30, 2020 with a mean time between preprint and journal publication of 65.4 days (range 0 to 271 days). The topics of the studies varied and included effects of clinical and public health interventions, associations of risk factors with COVID-19 symptoms, and ways to improve implementation of public health measures, such as social distancing. Almost a third of studies (21/67, 31%) were conducted in the United States, followed by Italy and Spain (n = 6, 9% each), and China (n = 5, 7%). The majority of studies reported public or non-profit funding sources (n=32, 49%) or that no funding was provided (n=24, 36%). Over half the studies also reported that the authors had no conflicts of interest (n=37, 53%).

### Discrepancies in study characteristics

Table 1 shows discrepancies in study characteristics reported in preprints and journal publications. The Table shows whether each study characteristic was reported or not; if a study characteristic was reported in both the preprint and journal publications, discrepancies in content are described. More preprints than journal publications reported funding source, author conflicts of interest and ethics approval; more journal publications than preprints reported participant demographics and study limitations. In all categories, most discrepancies occurred in the content of items that were reported, rather than in whether the item was present or not. For example, journal publications contained additional information on funding sources, conflicts of interest, demographic characteristics, and limitations, as well as more tables and figures compared to preprints (Table 1).

**Table 1:**
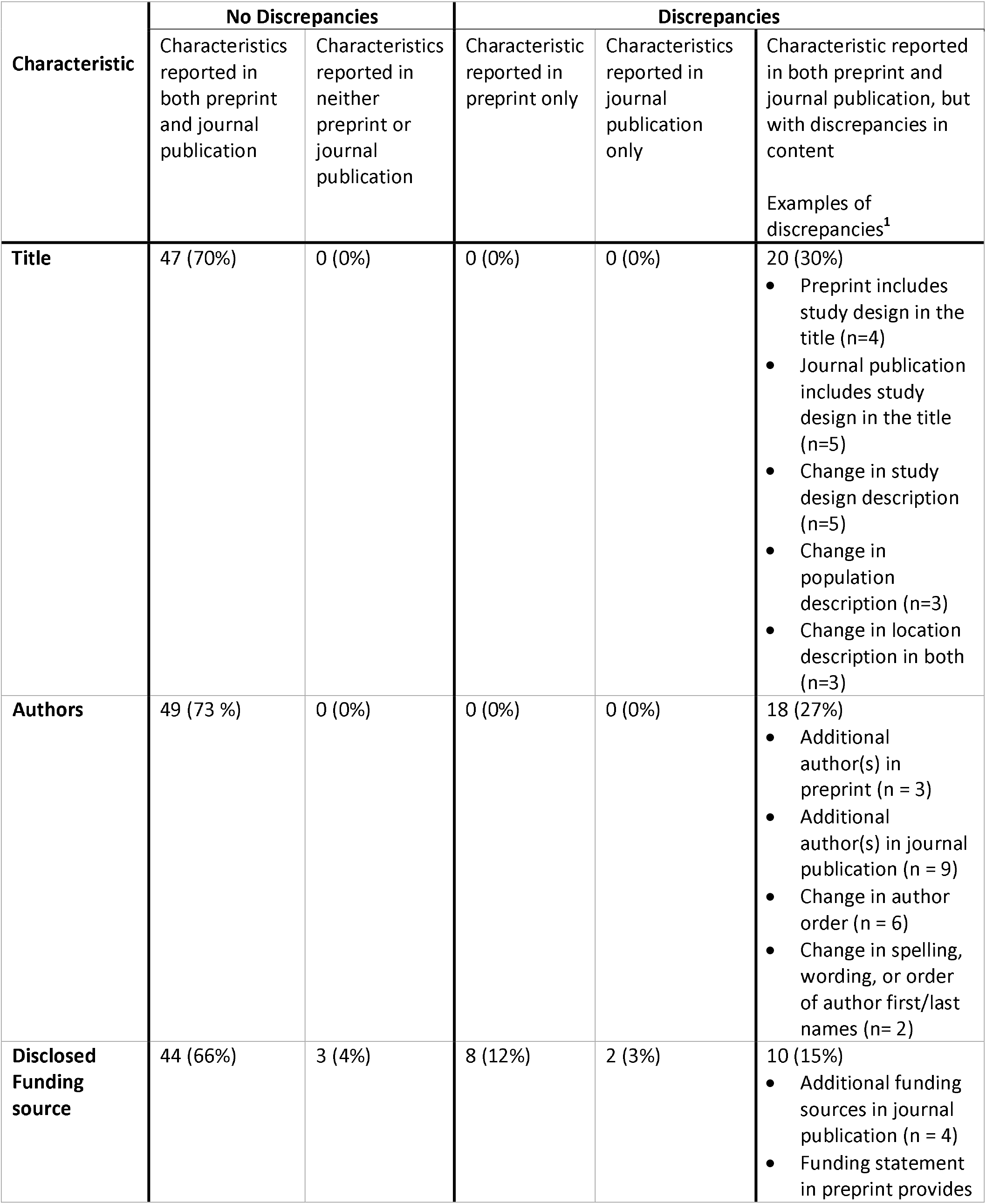
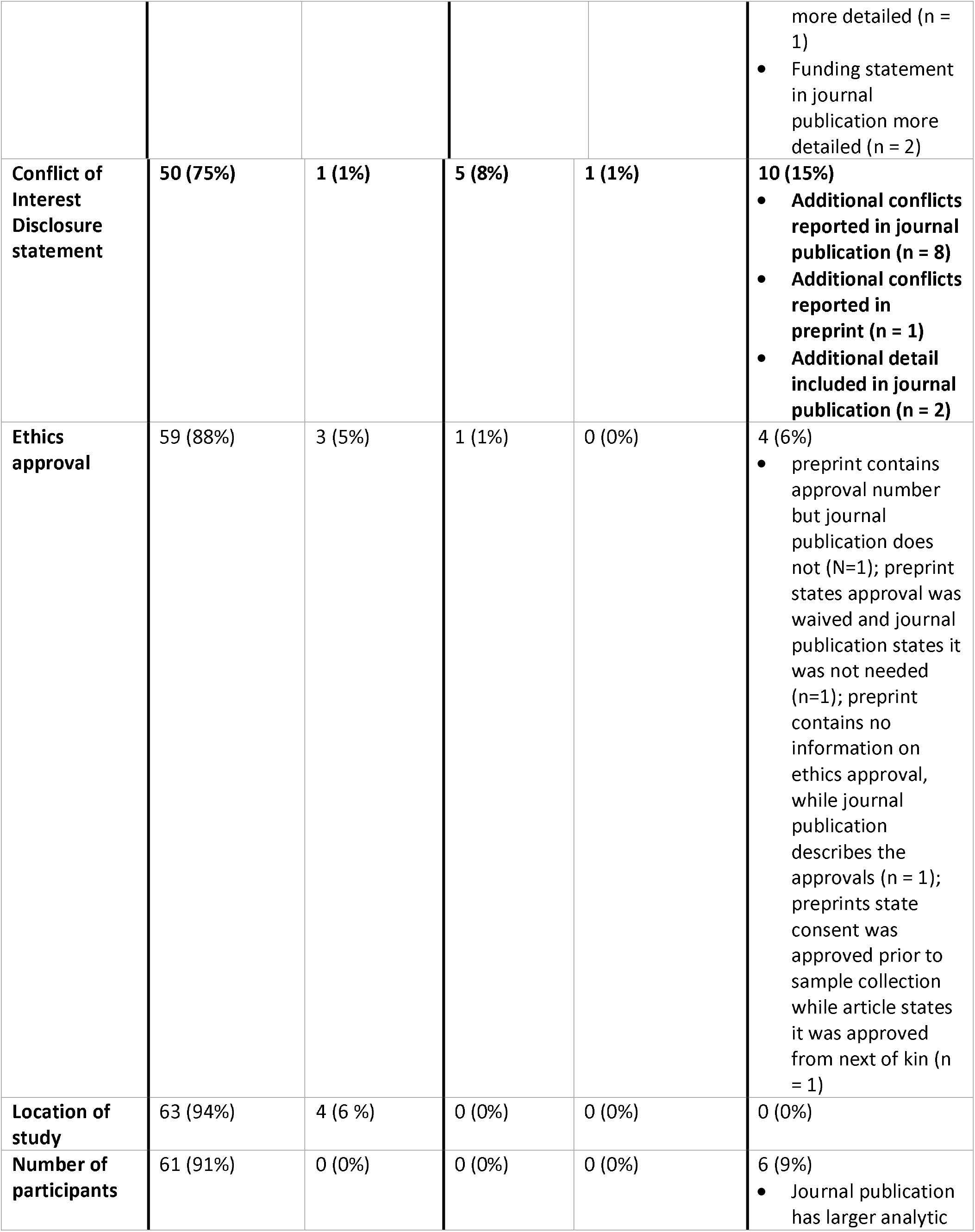
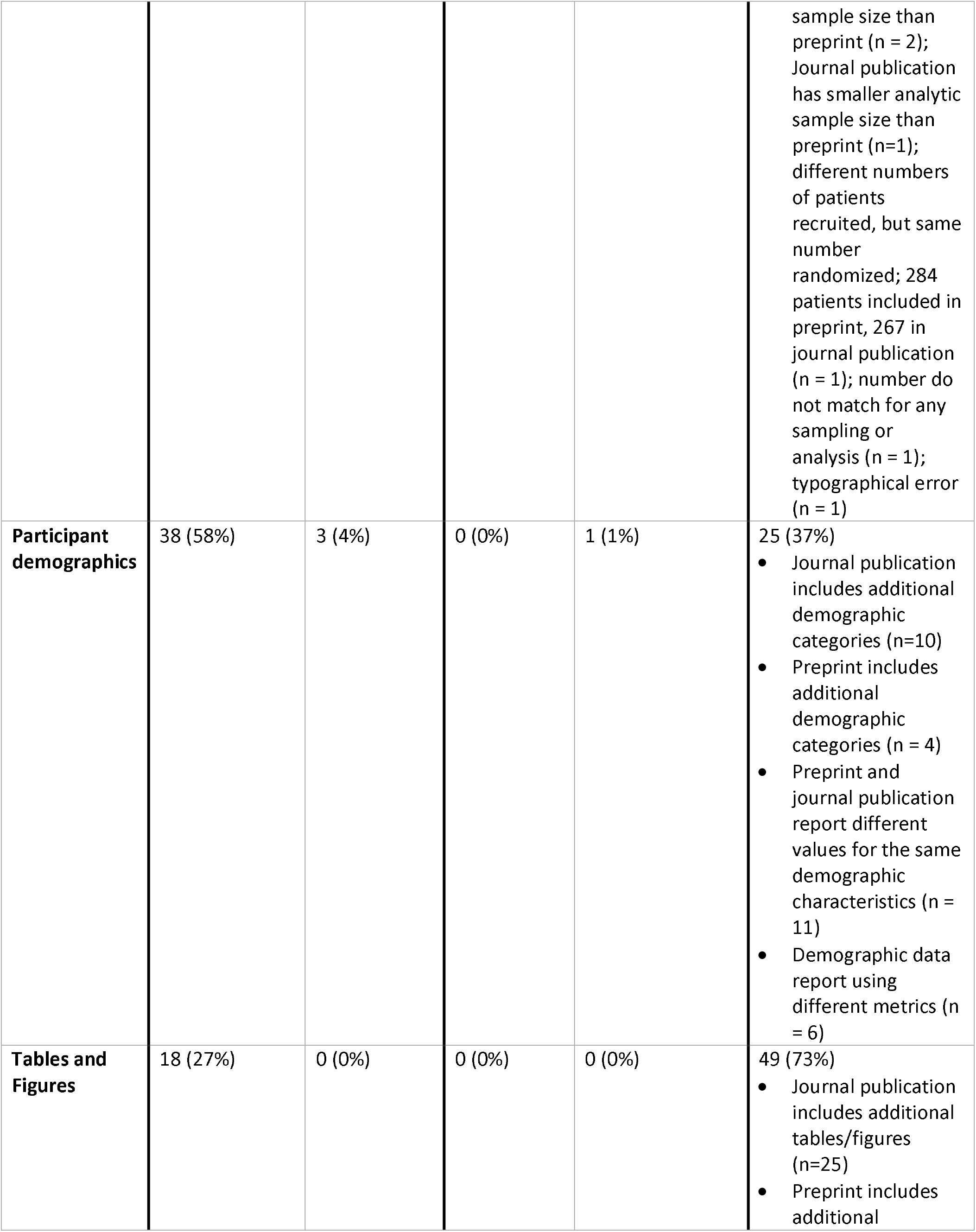
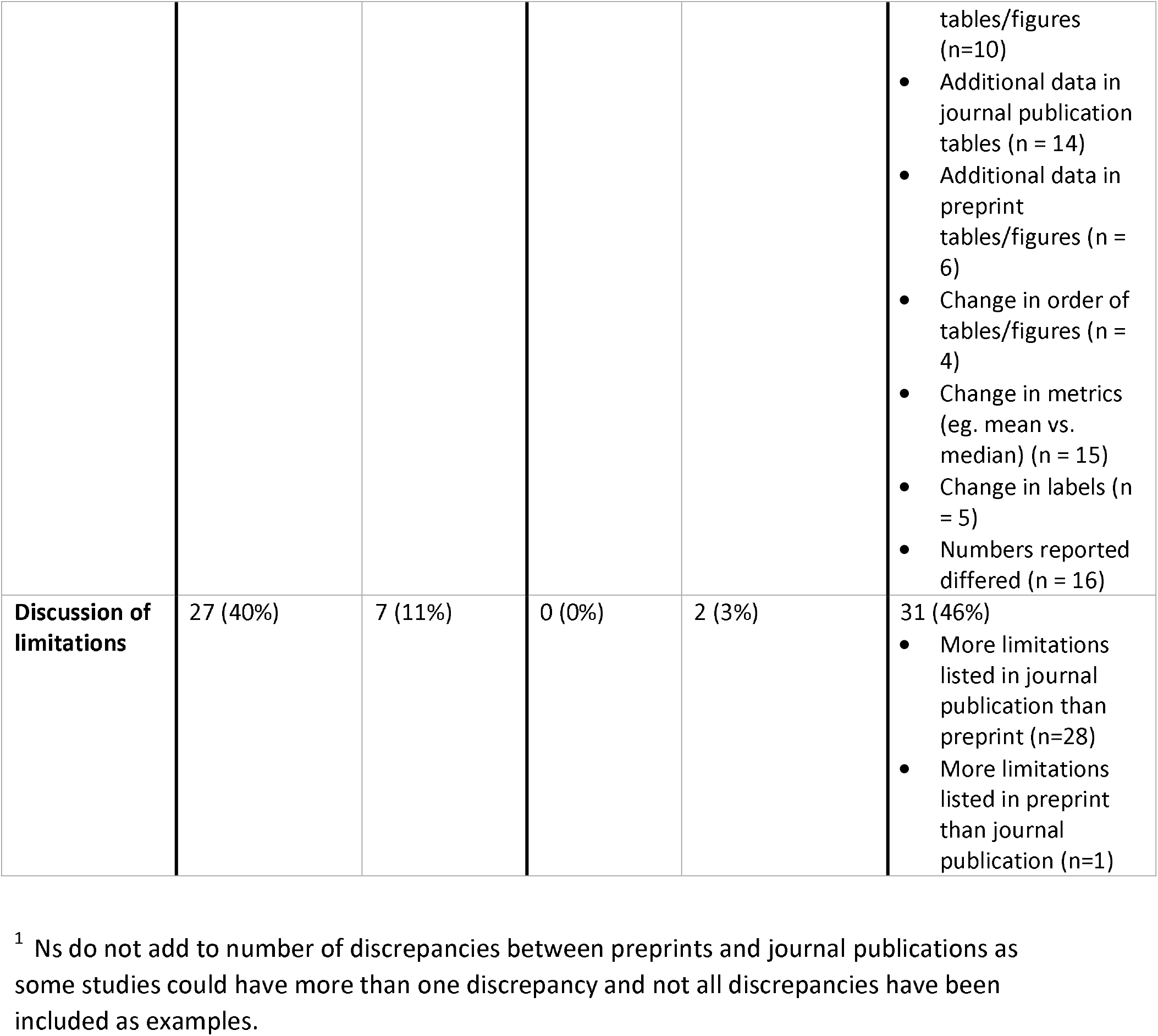
Discrepancies in Study Characteristics (n = 67 studies)

### Results reporting

Of the 67 studies, 23 (34%) had no discrepancies in results reporting between preprints and journal publications (Table 2). Twenty-three studies had outcomes that were missing from either the preprint or the journal publication. Fifteen (22%) studies had at least one outcome that was included in the journal publication, but not the preprint; 8 (12%) had at least one outcome that was reported in the preprint only. The included studies had multiple outcomes. The majority of studies with missing reported outcomes (16/23, 70%) had one outcome missing from either the preprint or journal publication. However, two studies had 5 outcomes missing from the journal publication, but reported in the preprint only.[22–25] As described in Table 2, these omissions included important clinical or harm outcomes. For example, one preprint omitted toxicity outcomes that were reported in the journal publication.[26,27]

**Table 2:** Discrepancies in Number of Outcomes Reported (N= 67 studies)

| **Type of Discrepancy** | **Number (%) of studies with at least 1 outcome that was reported only in the preprint or journal publication (n=67)** | **Number and description of outcomes across all studies that were reported only in the preprint or journal publication** |
| --- | --- | --- |
| **Outcome reported in journal publication only** | 15 (22%) | N = 19<br>1) Treatment-associated toxicities<br>2) Adverse reactions<br>3) Survival at ICU discharge<br>4) Creatine phosphokinase<br>5) Radiographic scale for acute respiratory distress syndrome<br>6) Time to negative swab<br>7) Time to RT-PCR negativity<br>8) Clinical outcomes at discharge<br>9) Ventilator status of those remaining hospitalized at end of follow up<br>10) Secondary composite - cardiovascular complications<br>11) Acute renal failure<br>12) Creatinine phosphokinase<br>13) Sequential organ failure assessment score<br>14) Length of stay<br>15) WHO Clinical Progression Scale<br>16) sCD14 levels related to corticoid treatment<br>17) Hospital Stay<br>18) Onset of symptoms<br>19) Mechanical ventilation or all-cause mortality at 21 days |
| **Outcome reported in preprint only** | 8 (12%) | N = 17<br>1) Oxygen support need<br>2) Invasive mechanical ventilation need<br>3) ICU need<br>4) Need for inotropics<br>5) Naso/oropharyngeal swab viral clearance<br>6) Final lymphocyte (cell/mm<sup>3</sup>)<br>7) Final CRP (mg/L)<br>8) Negative conversion of SARS-CoV-2 by 28 days<br>9) Negative conversion rate at 4-, 7-, 10-, 14- or |
| | | 21-day<br>10) Changes of CRP values and blood lymphocyte count<br>11) Rate of symptoms alleviation within 28-day<br>12) Safety endpoints<br>13) QTc ≥ 470 ms<br>14) Cumulative virus clearance rate vs different antiviral regimes in [a] all patients and [b] patients with moderate illness<br>15) Adverse events<br>16) Composite cardiovascular and renal failure<br>17) Nosocomial infections |

Table 3 shows the types of discrepancies in components of results reporting. We report the number of studies that had at least one discrepancy and, because studies have multiple outcomes, the number of discrepancies across all outcomes in the 67 studies. The most frequent types of discrepancies between outcomes reported in both preprints and journal publications were in the numerical values reported, statistical tests performed, subgroup analyses conducted, statistical significance reported, and timepoint at which the outcome was assessed (Table 3). The types of discrepancies were variable, although journal publications consistently included additional statistical analyses and subgroup analyses compared to preprints. Journal publications more frequently reported outcomes measured over a longer time period than preprints.

**Table 3:** Discrepancies in Components of Results Reporting for Outcomes Reported in Both Preprints and Journal Publications (N= 67 studies; 258 outcomes)

| **Type of Discrepancy** | **Number (%) of studies with at least 1 discrepancy between the preprint and journal publication (n=67)** | **Number (%) of outcomes across all studies that were discrepant between the preprint and journal publication (n=258)** | **Descriptive Examples<sup>1</sup>** |
| --- | --- | --- | --- |
| **Outcome measurement** | 6 (9%) | 8 (3%) | - Journal publication contains more detail on how outcome was measured compared to preprint (n=3)<br>- Journal publication reports an additional or different measurement than the one used for the same outcome in the preprint (e.g., preprint reports 4 adverse events, journal publication reports 12) (n=4) |
| **Units of measurement** | 3 (4%) | 3 (1%) | - e.g., journal publication reports events, total and percentage for mortality, preprint reports only percentage; median (IQR) reported in journal publication, mean (SD) in preprint |
| **Timepoint assessment was made** | 10 (15%) | 24 (9%) | - Journal publication reports outcomes measured over a longer timepoint than preprint (n=13)<br>- Journal publication reports additional interim time points compared to preprint (n=3) |
| **Numerical values reported** | 24 (36%) | 52 (20%) | - Differences in number of events or measurement values reported (n=17)<br>- Differences in numbers of participants or denominators (n = 5)<br>-<br>-More adverse events reported in journal publication than preprint (n = 4) |
| **Finding of statistical significance** | 11 (16%) | 16 (6%) | - Different p-value reported with no change in significance (n=3)<br>- Different p-value reported with change in significance; significant result reported in journal publication (n=1)<br>- In multivariate models, journal publication and preprint report different variables as being statistically significant (n=2) |
| **Statistical tests performed** | 17 (25%) | 31 (12%) | - Journal publication contains additional statistical analysis compared to preprint (n=7)<br>- Journal publication uses different statistical adjustments compared to preprint (n=7)<br>- Journal publication and preprint use different statistical tests for same data (n=3) |
| **Subgroup analyses conducted** | 14 (21%) | 24 (9%) | - Journal publication includes subgroup analysis not included in preprint (n=6)<br>- Journal publication finds statistically significant interaction for subgroup, preprint does not (n=1) |
| **Identifying the outcome as a primary or secondary outcome** | 1 (1%) | 3 (1%) | - e.g., preprint identifies the primary endpoint as safety; journal publication adds the secondary endpoint of exploration of efficacy |

<sup>1</sup> Ns do not add to number of reported discrepancies as some studies could have more than one discrepancy and not all discrepancies have been included as examples.

### Spin

At least one instance of spin occurred in the preprint, journal publication, or both in 30 (45%) of the 67 studies. Spin occurred in both preprints and journal publications in 23 / 67 (34%) studies, the preprint only in 5 (7%) studies, and the journal publications only in 2 (3%) studies (Table 4). Spin, in any category, was removed between the preprint and journal publication in 5 / 67(7%) studies; but added between the preprint and journal publication in 1 (1%) study.

**Table 4:**
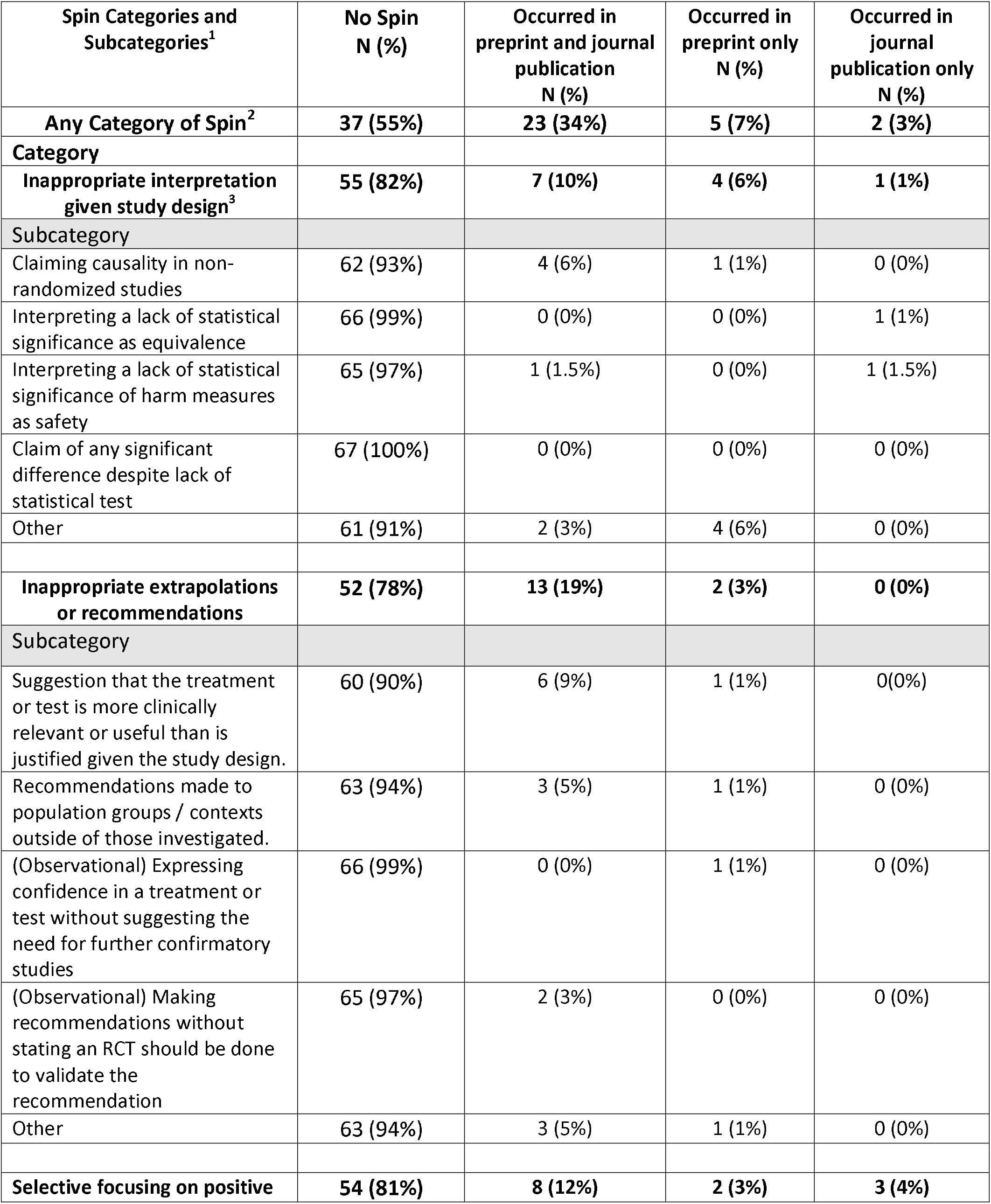
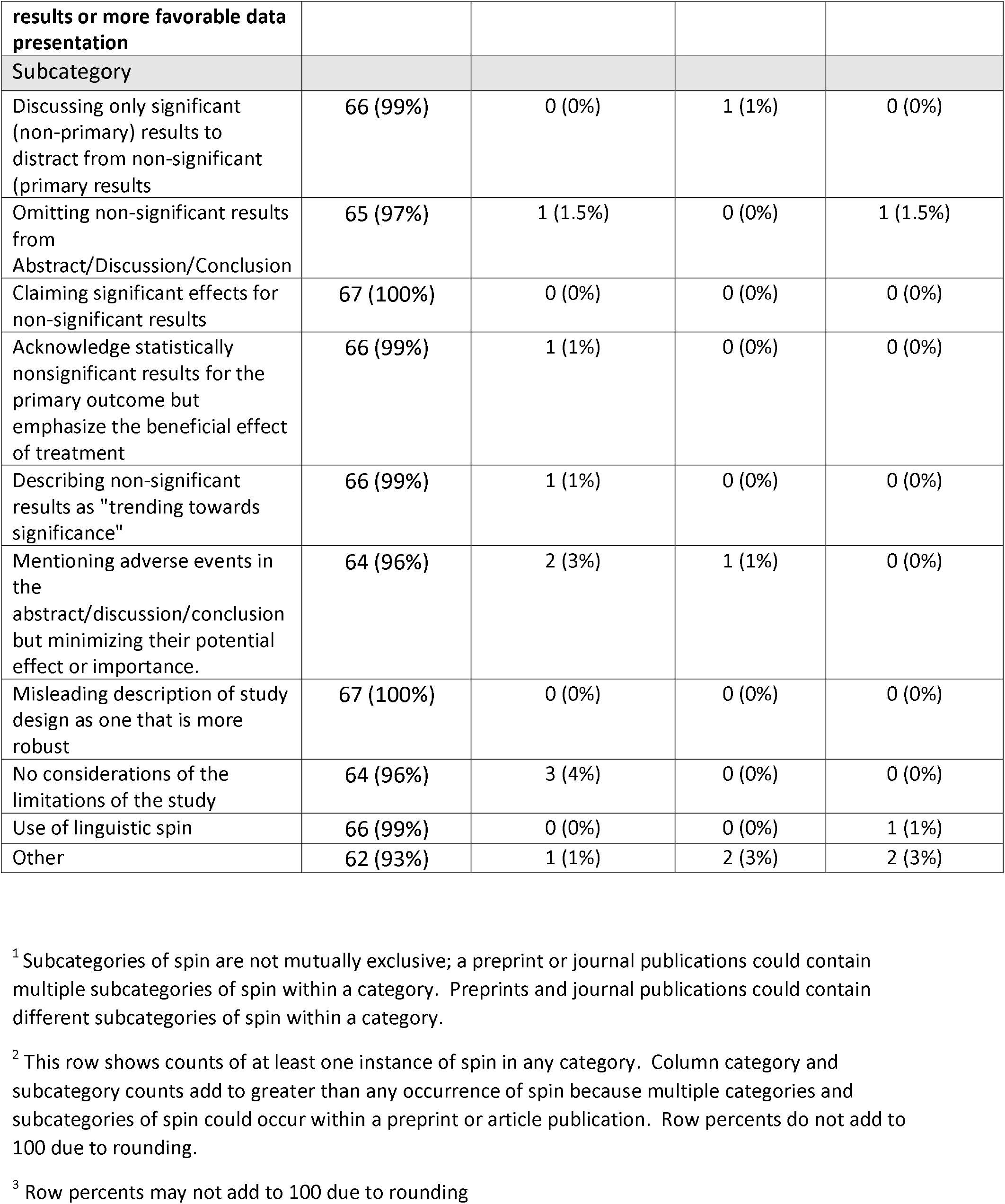
Categories of Spin in Preprints and Journal Publications (n = 67 studies)

Table 4 shows the categories of spin that occurred in preprints and their accompanying journal publications. Thirteen of 67 (19%) studies had changes in the type of spin present in the preprint versus the journal publication; 8 (12%) studies had at least one additional type of spin present in the preprint, 2 (3%) studies had at least one additional type of spin present in the journal publication. Inappropriate extrapolation or recommendations was the most frequently occurring type of spin in both preprints and journal publications (11/67, 16% of studies). This type of spin and inappropriate interpretation given the study design occurred more frequently in preprints than journal publications.

An example of inappropriate interpretation was found in both the preprint and journal publication for an open-label non-randomised trial: the study investigated the effect of hydroxychloroquine (and in combination with azithromycin) on SARS-Co-V-2 viral load. They found a statistically significant viral load reduction at day 6; however, despite the small sample size and non-randomised study design, they concluded that their findings were “so significant” and recommended that “COVID-19 patients be treated with hydroxychloroquine and azithromycin to cure their infection and to limit the transmission of the virus to other people in order to curb the spread of COVID-19 in the world.”[28,29] An example of inappropriate extrapolation or recommendations that occurred in both the preprint and journal publication is a study that recommended specific policy approaches that were not tested in the study: “The UK will shortly enter a new phase of the pandemic, in which extensive testing, contact tracing and isolation will be required to keep the spread of COVID-19. For this to succeed, adherence must be improved.”[30,31] This observational study aimed to identify factors associated with individuals’ adherence to self-isolation and lockdown measures; the authors did not aim to investigate public adherence to testing recommendations or contact tracing, nor test their efficacy.

### Sensitivity analysis

The mean time between preprint posting and journal article publication was 65.4 days (range 0 – 271) (Supplemental file, Table S1). No preprints were posted after the revision, acceptance or publication dates for the accompanying journal publication. One preprint was posted the same date as the publication date. Discrepancies in study characteristics, outcome reporting and spin changed minimally when the analyses were conducted after removing 7 studies where the preprint was posted up to 7 days before the revision, acceptance, or publication dates of the journal publication (Supplemental file, Tables S2 – S4).

## DISCUSSION

### Principal findings

Discrepancies between results reporting in preprints and their accompanying journal publications were frequent, but most often consisted of differences in content rather than a complete lack of reporting. Although infrequent, some outcomes that were not reported would have provided information that is critical for clinical decision making, such as clinical or harm outcomes that appeared only in the journal publication. The finding that outcomes reported in journal publications were measured over a longer time frame than outcomes reported in preprints indicates that the preprints were being used to publish preliminary or interim data. Preliminary or interim findings should be clearly labeled in preprints.

Although almost half of the preprints and journal publications contained spin, there was no clear difference in the types of spin. Spin is an enduring problem in the medical literature.[17] Our findings suggest that the identification and prevention of spin during journal peer review and editorial processes needs further improvement.

More preprints reported funding source, author conflicts of interest and ethics approval than journal publications. These differences may be due to the screening requirements of medRxiv, the main source of preprints in our sample. When reported in both, journal publications included more detailed information on funding source, conflicts of interest of authors, and demographics of the population studied. Journal publications also included more tables and figures, and more extensive discussion of limitations. Some of these differences may be due to more comprehensive reporting requirements of journals. Other changes, such as more information on the study population or greater discussion of limitations, may be due to requests for additional information during peer review.

Since preprints are posted without peer review and most journal publications in our sample were likely to be peer reviewed because they were identified from PubMed, our study indirectly investigates the impact of peer review on research articles. Articles may not have been peer-reviewed in similar ways. Authors may have made changes in their papers that were independent of peer review. We observed instances where peer review appeared to improve clarity (e.g, more detail on measurements)[32,33] or interpretation (e.g. requirement to present risk differences rather than just n (%) per treatment group).[34,35] Empirical evidence on the impact of peer review on manuscript quality is scarce. A study comparing submitted and published manuscripts found that the number of changes was relatively small and, similar to our study, primarily involved adding or clarifying information.[13] Some of the changes requested by peer reviewers were classified as having a negative impact on reporting, such as the addition of post-hoc subgroup analyses, statistical analyses that were not prespecified, or optimistic conclusions that did not reflect the trial results. In our sample, additions of subgroup and statistical analyses were common between preprints and journal publications, although we did not determine their appropriateness.

A small proportion of medRxiv preprints, 10% during the server’s first year, were published as journal publications.[5] Therefore, our sample could be limited to studies that their authors deemed of high enough quality to be eligible for submission to a journal. Or, our sample could be limited to articles that had not been rejected by a journal. It is possible that peer review was eliminating publications that were fundamentally unsound, while more quickly processing studies that were sound and useful. Under non-pandemic conditions, articles may undergo more revision. For example, peer reviewers may not suggest changes they think are less important, or editors may accept articles when they would have normally requested minor or major revisions. Thus, in this situation, peer review may mainly be playing the role of determining whether a study should be published in a journal or not.

There were minimal changes in the frequency and types of discrepancies between preprints and journal publications when we conducted a sensitivity analysis limiting our sample to studies where the preprints were published before the revision or acceptance date of the journal publication. This suggests that our findings are robust even when the sample is limited to preprints that could not have benefited from peer review. Given this finding and the observed similarities between preprints and their subsequent journal publications, our results suggest that peer review during the accelerated pace of COVID-19 research publication may not have provided much added value. The urgency related to dissemination of COVID-19 research could have led journals to fast-track publication by abbreviating editorial or peer review processes, resulting in fewer differences between preprints and journal publications.

### Comparison to other studies

Our results are consistent with other studies finding small changes in reporting between preprints and journal publications. A number of these studies have been limited by failing to assess the addition or deletion of outcomes and by the use of composite “scores” that included items related to risk of bias and reporting. In contrast to our study, in a matched sample of preprints and journal publications, Carnerio et al. found journal publications more likely to have conflict of interest statement than preprints. In a textual analysis using 5 different algorithms, Klein et al. found very little difference in text between preprints and articles in a large matched sample.[9] We also noted preprints and journal publications that were almost identical, or had very minor differences such as corrections of typos. Other studies are limited by comparing unmatched samples of preprints and articles. In a comparison of 13 preprints and 16 articles on COVID-19 that were not reporting on the same studies, Kataoka et al. found no significant differences in risk of bias or spin in titles and conclusions.[11]

We found similar changes in numerical results to Oikonomidi et al. who compared 66 preprint-article pairs for COVID-19 studies and found 25 (38%) of studies had changes.[12] Oikonomidi classified 16 of these changes as “important” based on 1) an increase or decrease by ≥ 10% of the initial value in any effect estimate and/or 2) a change in the p-value crossing the threshold of 0.05, for any study outcome. We did not classify changes based on magnitude or threshold p-values because changes in numerical values may be related to other components of outcome reporting that we observed, such as changes to follow-up times or the use of different statistical tests. Furthermore, deviations from a p-value of 0.05 do not necessarily indicate changes in scientific or clinical significance. We examined changes in multiple components of outcome reporting that are considered essential, not just the numerical value of the outcome.[16,21] The diversity of studies included in our sample would make any categorizations of scientific or clinical significance difficult and subjective. For example, studies were observational and experimental and not all studies conducted statistical analysis. The topics of the studies included tests of clinical and public health interventions, associations of risk factors with COVID-19 symptoms, and ways to improve implementation of public health measures, such as social distancing.

### Strengths and weaknesses of this study

We selected studies from the Cochrane COVID-19 Register rather than conducting a literature search. However, as the Cochrane COVID-19 Register has been optimized to identify COVID-19 clinical research for systematic reviews, we feel the search was comprehensive for identifying COVID-19 studies related to treatment or prevention that are most likely to have an impact on clinical practice or health policy. As a study-based register, all records related to a study are identified, enabling us to obtain all preprint and journal publication versions for a single study. Second, we compared the first version of the preprint with the final journal publication. We may have identified a different number of discrepancies if we compared later versions of the preprint with the journal publication. Third, although clinically important, our focus on COVID-19 research may not be representative of other types of research published as preprints, then journal publications. This study should be replicated in a sample of non-COVID related interventional and observational clinical studies. Future research could also include assessment of outcome reporting components and spin in preprints that have not been published in journals. Fourth, although we compared non-peer-reviewed preprints to their accompanying journal publications, we did not directly assess the effects of peer review. Finally, coders were not blinded to the source or authors of preprints and journal publications as this was not feasible and there is no evidence that it would alter the decisions made.

## CONCLUSIONS

The COVID-19 preprints and their subsequent journal publications were largely similar in reporting of study characteristics, outcomes and spin in interpretation. However, given the urgent need for valid and reliable research on COVID-19 treatment and prevention, even a few important discrepancies could impact decision making. All COVID-19 studies, whether published as preprints or journal publications, should be critically evaluated for discrepancies in outcome reporting or spin, such as failure to report data on harms or overly optimistic conclusions.

## Data Availability

Data from this study is available in OSF project file (https://osf.io/5ru8w/?view_only=fe509bf54c104354a1e12f011bdff66a).

## Data access

LB had full access to all the data in the study and takes responsibility for the integrity of the data and the accuracy of the data analysis.

## Author contributions

LB conceived the project. All authors made substantial contributions to design of the work, the acquisition of data, analysis, and interpretation of data for the work. LB drafted the paper and all authors revised it critically for important intellectual content. All authors have approved the final manuscript. LB serves as guarantor for all aspects of the work.

Transparency declaration: LB affirms that the manuscript is an honest, accurate, and transparent account of the study being reported; that no important aspects of the study have been omitted; and that any discrepancies from the study as planned have been explained.

## REFERENCES

1 All that’s fit to preprint. Nat Biotechnol 2020;38:507–507. doi:10.1038/s41587-020-0536-x

2 Flanagin A, Fontanarosa PB, Bauchner H. Preprints involving medical research: do the benefits outweigh the challenges? JAMA 2020;324:1840. doi:10.1001/jama.2020.20674

3 Jung YE (Grace), Sun Y, Schluger NW. Effect and reach of medical articles posted on preprint servers during the COVID-19 pandemic. JAMA Intern Med Published Online First: 9 November 2020. doi:10.1001/jamainternmed.2020.6629

4 Kirkham JJ, Penfold NC, Murphy F, et al. Systematic examination of preprint platforms for use in the medical and biomedical sciences setting. BMJ Open 2020;10:e041849. doi:10.1136/bmjopen-2020-041849

5 Krumholz HM, Bloom T, Sever R, et al. Submissions and downloads of preprints in the first year of medRxiv. JAMA 2020;324:1903–5. doi:10.1001/jama.2020.17529

6 Krumholz HM, Bloom T, Ross J. Preprints can fill a void in times of rapidly changing science. STAT. 2020.https://www.statnews.com/2020/01/31/preprints-fill-void-rapidly-changing-science/ (accessed 17 Dec 2020).

7 Bloom T. Shepherding preprints through a pandemic. BMJ 2020;371:m4703. doi:10.1136/bmj.m4703

8 Carneiro CFD, Queiroz VGS, Moulin TC, et al. Comparing quality of reporting between preprints and peer-reviewed articles in the biomedical literature. Res Integr Peer Rev 2020;5:16. doi:10.1186/s41073-020-00101-3

9 Klein M, Broadwell P, Farb SE, et al. Comparing published scientific journal articles to their pre-print versions. In: Proceedings of the 16th ACM/IEEE-CS on Joint Conference on Digital Libraries. New York, NY, USA: : Association for Computing Machinery 2016. 153–62. doi:10.1145/2910896.2910909

10 Nicolalde B, Añazco D, Mushtaq M, et al. Citations and publication rate of preprints on pharmacological interventions for COVID-19: the good, the bad and, the ugly. Version 2. Res Sq Published Online First: 8 September 2020. doi:10.21203/rs.3.rs-34689/v2

11 Kataoka Y, Oide S, Ariie T, et al. COVID-19 randomized controlled trials in medRxiv and PubMed. Eur J Intern Med 2020;81:97–9. doi:10.1016/j.ejim.2020.09.019

12 Oikonomidi T, Boutron I, Pierre O, et al. Changes in evidence for studies assessing interventions for COVID-19 reported in preprints: meta-research study. BMC Med 2020;18:402. doi:10.1186/s12916-020-01880-8

13 Hopewell S, Collins GS, Boutron I, et al. Impact of peer review on reports of randomised trials published in open peer review journals: retrospective before and after study. BMJ 2014;349:g4145. doi:10.1136/bmj.g4145

14 Lazarus C, Haneef R, Ravaud P, et al. Peer reviewers identified spin in manuscripts of nonrandomized studies assessing therapeutic interventions, but their impact on spin in abstract conclusions was limited. J Clin Epidemiol 2016;77:44–51. doi:10.1016/j.jclinepi.2016.04.012

15 Chan A-W, Hróbjartsson A, Haahr MT, et al. Empirical evidence for selective reporting of outcomes in randomized trials: comparison of protocols to published articles. JAMA 2004;291:2457. doi:10.1001/jama.291.20.2457

16 Mathieu S, Boutron I, Moher D, et al. Comparison of registered and published primary outcomes in randomized controlled trials. JAMA 2009;302:977–84. doi:10.1001/jama.2009.1242

17 Chiu K, Grundy Q, Bero L. ‘Spin’ in published biomedical literature: A methodological systematic review. PLoS Biol 2017;15:e2002173. doi:10.1371/journal.pbio.2002173

18 Boutron I, Ravaud P. Misrepresentation and distortion of research in biomedical literature. Proc Natl Acad Sci 2018;115:2613–9. doi:10.1073/pnas.1710755115

19 Bero L, Lawrence R, Leslie L, et al. Comparison of preprints with peer-reviewed publications on COVID-19: discrepancies in results reporting and conclusions. 2020.https://osf.io/j62eu (accessed 17 Dec 2020).

20 Harris PA, Taylor R, Thielke R, et al. Research electronic data capture (REDCap): a metadata-driven methodology and workflow process for providing translational research informatics support. J Biomed Inform 2009;42:377–81. doi:10.1016/j.jbi.2008.08.010

21 Zarin DA, Tse T, Williams RJ, et al. The ClinicalTrials.gov Results Database: update and key Issues. N Engl J Med 2011;364:852–60. doi:10.1056/NEJMsa1012065

22 Borba MGS, Val FFA, Sampaio VS, et al. Effect of High vs Low Doses of Chloroquine Diphosphate as Adjunctive Therapy for Patients Hospitalized With Severe Acute Respiratory Syndrome Coronavirus 2 (SARS-CoV-2) Infection: A Randomized Clinical Trial. JAMA Netw Open 2020;3:e208857. doi:10.1001/jamanetworkopen.2020.8857

23 Borba MGS, Val FFA, Sampaio VS, et al. Chloroquine diphosphate in two different dosages as adjunctive therapy of hospitalized patients with severe respiratory syndrome in the context of coronavirus (SARS-CoV-2) infection: Preliminary safety results of a randomized, double-blinded, phase IIb clinical trial (CloroCovid-19 Study). Version 1. MedRxiv Prepr 2020;:2020.04.07.20056424. doi:10.1101/2020.04.07.20056424

24 Tang W, Cao Z, Han M, et al. Hydroxychloroquine in patients with COVID-19: an open-label, randomized, controlled trial. Version 1. MedRxiv Prepr 2020;:2020.04.10.20060558. doi:10.1101/2020.04.10.20060558

25 Tang W, Cao Z, Han M, et al. Hydroxychloroquine in patients with mainly mild to moderate coronavirus disease 2019: open label, randomised controlled trial. BMJ 2020;369:m1849. doi:10.1136/bmj.m1849

26 Weber AG, Chau AS, Egeblad M, et al. Nebulized in-line endotracheal dornase alfa and albuterol administered to mechanically ventilated COVID-19 patients: a case series. MedRxiv Prepr Published Online First: 15 May 2020. doi:10.1101/2020.05.13.20087734

27 Weber AG, Chau AS, Egeblad M, et al. Nebulized in-line endotracheal dornase alfa and albuterol administered to mechanically ventilated COVID-19 patients: a case series. Mol Med 2020;26:91. doi:10.1186/s10020-020-00215-w

28 Gautret P, Lagier J-C, Parola P, et al. Hydroxychloroquine and azithromycin as a treatment of COVID-19: results of an open-label non-randomized clinical trial. medRxiv 2020;:2020.03.16.20037135. doi:10.1101/2020.03.16.20037135

29 Gautret P, Lagier J-C, Parola P, et al. Hydroxychloroquine and azithromycin as a treatment of COVID-19: results of an open-label non-randomized clinical trial. Int J Antimicrob Agents 2020;56:105949. doi:10.1016/j.ijantimicag.2020.105949

30 Smith LE, Amlôt R, Lambert H, et al. Factors associated with adherence to self-isolation and lockdown measures in the UK; a cross-sectional survey. MedRxiv Prepr 2020;:2020.06.01.20119040. doi:10.1101/2020.06.01.20119040

31 Smith LE, Amlt R, Lambert H, et al. Factors associated with adherence to self-isolation and lockdown measures in the UK: a cross-sectional survey. Public Health 2020;187:41–52. doi:10.1016/j.puhe.2020.07.024

32 Chorin E, Wadhwani L, Magnani S, et al. QT interval prolongation and torsade de pointes in patients with COVID-19 treated with hydroxychloroquine/azithromycin. Heart Rhythm 2020;17:1425–33. doi:10.1016/j.hrthm.2020.05.014

33 Chorin E, Wadhwani L, Magnani S, et al. QT Interval Prolongation and Torsade De Pointes in Patients with COVID-19 treated with Hydroxychloroquine/Azithromycin. MedRxiv Prepr 2020;:2020.04.27.20074583. doi:10.1101/2020.04.27.20074583

34 Agarwal A, Mukherjee A, Kumar G, et al. Convalescent plasma in the management of moderate COVID-19 in India: an open-label parallel-arm phase II multicentre randomized controlled trial (PLACID Trial). Version 1. MedRxiv Prepr 2020;:2020.09.03.20187252. doi:10.1101/2020.09.03.20187252

35 Agarwal A, Mukherjee A, Kumar G, et al. Convalescent plasma in the management of moderate covid-19 in adults in India: open label phase II multicentre randomised controlled trial (PLACID Trial). BMJ 2020;371:m3939. doi:10.1136/bmj.m3939

