## Supplementary material for "Comparison of preprints and final journal publications from COVID-19 Studies: Discrepancies in results reporting and spin in interpretation": Tables S1-S4

**Supplemental Files**

**Table S1: Timing of preprint to journal publication (days)**

**Table S2: Sensitivity Analysis of Discrepancies in Study Characteristics**

**Table S3: Sensitivity Analysis of Discrepancies in Outcome Reporting**

**Table S4: Sensitivity Analysis of Categories of Spin in Preprints and Journal Publications**

**Table S1: Timing of preprint to journal publication (days)**

|  | **Days from preprint to published, mean (range)** |
| --- | --- |
| **All Studies (n=67)** | 65.4 (0 - 271) |
| **Subgroup: Preprint posted before submission to journal (n=32)** | 87.1 (10 - 271) |
| **Subgroup: Preprint posted after submission to journal (n=27)** | 52.2 (0 - 120) |

**Table S2: Sensitivity Analysis of Discrepancies in Study Characteristics (n=60) ^a^**

|  | **No Discrepancies** | | **Discrepancies** | | |
| --- | --- | --- | --- | --- | --- |
|  | **Reported in Both, No. (%)** | **Reported in Neither, No. (%)** | **Reported in Both With Discrepancies, No. (%)** | **Reported in Preprint Only, No. (%)** | **Reported in Journal Publication Only, No. (%)** |
| **Title** | 44 (73) | 0 (0) | 16 (27) | 0 (0) | 0 (0) |
| **Authors** | 43 (72) | 0 (0) | 17 (28) | 0 (0) | 0 (0) |
| **Disclosed Funding Source** | 39 (65) | 3 (5) | 10 (17) | 6 (10) | 2 (3) |
| **COI Disclosure Statement** | 45 (75) | 1 (2) | 9 (15) | 4 (7) | 1 (2) |
| **Ethics Approval** | 54 (90) | 2 (3) | 4 (7) | 0 (0) | 0 (0) |
| **Location of Study** | 56 (93) | 4 (7) | 0 (0) | 0 (0) | 0 (0) |
| **Number of Participants** | 54 (90) | 0 (0) | 6 (10) | 0 (0) | 0 (0) |
| **Participant Demographics** | 34 (57) | 3 (5) | 22 (37) | 0 (0) | 1 (2) |
| **Tables and Figures** | 15 (25) | 0 (0) | 45 (75) | 0 (0) | 0 (0) |
| **Discussion of Limitations** | 23 (38) | 6 (10) | 30 (50) | 0 (0) | 1 (2) |

^a^ Studies that had a preprint posted on-or-after the date of revision, acceptance, or publication were removed. This removed 1 study. Due to differences in journal reporting of these dates, there was overlap in those studies and no comparison in others. Therefore, we expanded the studies removed to include those with preprints posted 1-7 days before the date of revision, acceptance, or publication, thus removing 7 studies from the sensitivity analysis.

**Table S3: Sensitivity Analysis of Discrepancies in Outcome Reporting (n=60) ^a^**

|  | **Number (%) of studies with at least 1 discrepancy**  **n=60** | **Number (%) of Outcomes n=242** |
| --- | --- | --- |
| **Outcome in journal publication only** | 14 (23) | 18 (7) |
| **Outcome in preprint only** | 7 (12) | 16 (7) |
| **Outcome measurement** | 5 (8) | 7 (3) |
| **Units of measurement** | 3 (5) | 3 (1) |
| **Timepoint assessment was made** | 10 (17) | 24 (10) |
| **Numerical values reported** | 23 (38) | 49 (20) |
| **Finding of statistical significance** | 11 (18) | 16 (7) |
| **Statistical tests performed** | 16 (27) | 30 (12) |
| **Subgroup analyses conducted** | 13 (22) | 23 (10) |
| **Identifying the outcome as a primary or secondary outcome** | 1 (2) | 3 (1) |

^a^ Studies that had a preprint posted on-or-after the date of revision, acceptance, or publication were removed. This removed 1 study. Due to differences in journal reporting of these dates, there was overlap in those studies and no comparison in others. Therefore, we expanded the studies removed to include those with preprints posted 1-7 days before the date of revision, acceptance, or publication, thus removing 7 studies from the sensitivity analysis.

**Table S4: Sensitivity Analysis of Categories of Spin in Preprints and Journal Publications (n=60) ^a^**

|  | **Neither, No. (%)** | **Both, No. (%)** | **Preprint Only, No. (%)** | **Journal Publication Only, No. (%)** |
| --- | --- | --- | --- | --- |
| **Inappropriate interpretation given study design** | **49 (82)** | **6 (10)** | **4 (7)** | **1 (2)** |
| Claiming causality in non-randomized studies | 56 (93) | 3 (5) | 1 (2) | 0 (0) |
| Interpreting a lack of statistical significance as equivalence | 59 (98) | 0 (0) | 0 (0) | 1 (2) |
| Interpreting a lack of statistical significance of harm measures as safety | 58 (97) | 1 (2) | 0 (0) | 1 (2) |
| Claim of any significant difference despite lack of statistical test | 60 (100) | 0 (0) | 0 (0) | 0 (0) |
| Other | 54 (90) | 2 (3) | 4 (7) | 0 (0) |
| **Inappropriate extrapolations or recommendations** | **46 (77)** | **12 (20)** | **2 (3)** | **0 (0)** |
| Suggestion that the treatment or test is more clinically relevant or useful than is justified given the study design. | 54 (90) | 5 (8) | 1 (2) | 0 (0) |
| Recommendations made to population groups / contexts outside of those investigated. | 56 (93) | 3 (5) | 1 (2) | 0 (0) |
| (Observational) Expressing confidence in a treatment or test without suggesting the need for further confirmatory studies | 59 (98) | 0 (0) | 1 (2) | 0 (0) |
| (Observational) Making recommendations without stating an RCT should be done to validate the recommendation | 59 (98) | 1 (2) | 0 (0) | 0 (0) |
| Other | 56 (93) | 3 (5) | 1 (2) | 0 (0) |
| **Selective focusing on positive results or more favorable data presentation** | **48 (80)** | **7 (12)** | **2 (3)** | **3 (5)** |
| Discussing only significant (non-primary) results to distract from non-significant (primary results | 59 (98) | 0 (0) | 1 (2) | 0 (0) |
| Omitting non-significant results from Abstract/Discussion/Conclusion | 58 (97) | 1 (2) | 0 (0) | 1 (2) |
| Claiming significant effects for non-significant results | 60 (100) | 0 (0) | 0 (0) | 0 (0) |
| Acknowledge statistically nonsignificant results for the primary outcome but emphasize the beneficial effect of treatment | 59 (98) | 1 (2) | 0 (0) | 0 (0) |
| Describing non-significant results as "trending towards significance" | 59 (98) | 1 (2) | 0 (0) | 0 (0) |
| Mentioning adverse events in the abstract/discussion/conclusion but minimizing their potential effect or importance. | 58 (97) | 1 (2) | 1 (2) | 0 (0) |
| Misleading description of study design as one that is more robust | 60 (100) | 0 (0) | 0 (0) | 0 (0) |
| No considerations of the limitations of the study | 58 (97) | 2 (3) | 0 (0) | 0 (0) |
| Use of linguistic spin | 59 (98) | 0 (0) | 0 (0) | 1 (2) |
| Other | 55 (92) | 1 (2) | 2 (3) | 2 (3) |

^a^ Studies that had a preprint posted on-or-after the date of revision, acceptance, or publication were removed. This removed 1 study. Due to differences in journal reporting of these dates, there was overlap in those studies and no comparison in others. Therefore, we expanded the studies removed to include those with preprints posted 1-7 days before the date of revision, acceptance, or publication, thus removing 7 studies from the analysis
